# Quantifying anti-trypanosomal treatment effects in chronic indeterminate Chagas disease: an individual patient data meta-analysis of two proof of concept trials

**DOI:** 10.1101/2024.07.14.24310398

**Authors:** James A Watson, Cintia Cruz, Fabiana Barreira, Colin Forsyth, Alejandro Schijman, Rhys Peploe, Frauke Assmus, Caitlin Naylor, Jennifer Lee, Somya Mehra, Joel Tarning, Faustino Torrico, Joaquim Gascon, Lourdes Ortiz, Isabela Ribeiro, Sergio Sosa-Estani, Craig Tipple, Stéphane Hugonnet, Philippe J Guérin, Laurent Fraisse, Maria-Jesus Pinazo, Nicholas J White, the E1224 and BENDITA study groups

## Abstract

**Background:** The current antiparasitic treatment for chronic Chagas disease of 8 weeks daily benznidazole or nifurtimox is poorly tolerated and reaches only a small minority of those with chronic infections. Defining parasitological cure is compromised by the low blood trypomastigote densities, which fluctuate close to or below the limit of qPCR detection.

**Methods:** To address this limitation and improve the assessment of parasitological cure we developed a probabilistic model of therapeutic efficacy based on serial qPCR data. We pooled clinical and laboratory data from two prospective trials in Bolivian adults with chronic indeterminate Chagas disease. In both trials randomised arms included placebo or standard of care benznidazole (300mg/day for 8 weeks). In the first trial, the experimental arms were fosravuconazole monotherapies (400mg/week for 4 or 8 weeks, or 200mg/week for 8 weeks); in the second trial the experimental arms were shorter or lower dose benznidazole regimens (300mg/day for 2 or 4, or 150mg/day for 4 weeks), or combinations of fosravuconazole 300mg weekly for 8 weeks with either benznidazole 150mg/day for 4 weeks or benznidazole 300mg/week for 8 weeks. Serial parasite densities were estimated from triplicate qPCRs targeting *T. cruzi* satellite DNA taken from one to three 5 or 10ml blood samples at 8-12 visits over one year. Treatment efficacies were estimated under a hierarchical Bayesian model, taking as input serial cycle threshold (Ct) data grouped by time point, blood draw and technical replicate. The primary analysis was done in a per-protocol population defined as patients randomised to placebo or patients who took an active treatment >80% of the allocated treatment duration.

**Results:** The two trials randomised 441 patients. 34,804 qPCR Ct values were recorded over 5,402 unique visits, comprising 449 participant years follow-up. In a per-protocol population (*n*=424), an estimated 81% (95% Credible Interval [CrI] 70 to 89%, *n*=69) had parasitological cure following benznidazole 300mg/day for 8 weeks. All other benznidazole regimens had similar estimated cure proportions (95% CrIs >63%) except the 2-week regimen (63% cured [95%CrI 43-81%], *n*=27, probability of inferiority relative 8-week: 0.95). Recurrent parasitaemias following benznidazole were at substantially lower densities than at baseline. In comparison, only 3.9% of patients allocated to placebo were cured (95%CrI 1 to 9%, *n*=77). Fosravuconazole was relatively ineffective: 23% cured following 400mg for 8 weeks (95%CrI, 10 to 40%, *n*=45); 9% following 400mg for 4 weeks (95%CrI 3 to 21%, *n*=46); and 2% following 200mg for 8 weeks (95%CrI 0 to 11%, *n*=48). Recurrent parasitaemias one year after fosravuconazole treatment were only slightly lower than at baseline. Fosravuconazole caused dose-dependent increases in liver transaminases.

**Conclusions:** Therapeutic assessments in Chagas disease must account probabilistically for qPCR test performance and low density post treatment parasitaemias. In Bolivian chronic Chagas disease, weekly dosing for eight weeks or daily dosing over four weeks both appear as effective as the current eight weeks daily regimen. The total dose of benznidazole in the current standard of care regimen is excessive.

## Background

Chronic infection with the parasite *T. cruzi* is characterised by very low circulating trypomastigote densities in blood (usually less than 1 parasite per ml). The trypanosome parasites cycle repeatedly between the blood (as motile trypomastigotes) and the tissues where mitotic cell division of amastigotes takes place [1]. After an initial proliferative phase, parasite multiplication is held in check by the host immune system, resulting in a quasi-steady state evidenced by stable trypomastigote densities in blood [2]. Several decades later, the cumulative tissue damage resulting from the amastigote stage infection and the host response causes organ dysfunction (particularly affecting the heart or gastrointestinal tract) in around one-third of people living with chronic Chagas disease [1, 3]. There are currently only two available antiparasitic treatments for chronic *T. cruzi* infection, nifurtimox and benznidazole, both of which were introduced over fifty years ago. The Pan American Health Organisation recommends trypanocidal treatment for adult patients with chronic *T. cruzi* infection and no specific organ damage (chronic indeterminate Chagas: conditional recommendation, based on low certainty regarding the effects of the intervention) [4].

The currently recommended eight-week nifurtimox or benznidazole treatment regimens are not well tolerated by adults, and so there are high rates of patient discontinuation [5–7]. WHO has put 75% treatment coverage in Chagas disease as one of its neglected tropical diseases roadmap objectives, and antiparasitic treatment is likely to be highly cost-effective [8], but still today most people with chronic *T. cruzi* infection do not receive anti-trypanosomal treatment. This dismal situation results from many factors including poor access to treatment and lack of available diagnosis [4].

Benznidazole is the more widely used drug for Chagas disease, although the optimum dosage and duration of treatment, drug metabolism, and mechanisms of action and toxicity are all still not well characterised. To improve tolerability, and thus increase access to effective treatment, shorter and intermittent course benznidazole regimens have been investigated. The Drugs for Neglected Diseases *initiative* (DND*i* ) sponsored two recent studies in chronic indeterminate Chagas disease. The first (ClinicalTrials.gov, NCT01489228) evaluated the antiparasitic effect of the antifungal azole drug fosravuconazole (E1224), taken at two different weekly doses for 4 or 8 weeks as a monotherapy [6]. The second, BENDITA (ClinicalTrials.gov, NCT03378661), evaluated several short-course benznidazole regimens, some in combination with fosravuconazole, and one intermittent weekly regimen in combination with fosravuconazole [7]. Both these studies had very similar designs and were done in similar patient populations in the same clinical sites in Bolivia. Assessing antiparasitic therapeutics in chronic indeterminate Chagas disease poses several important challenges. First, clinically significant pathology does not usually occur for decades after primary infection and bears an uncertain relationship to parasite burden. Second, trypomastigote densities in blood are extremely low (often below the limit of accurate detection) and third, seroreversion is often slow in cured individuals [9]. Nearly all recent studies of antiparasitic drugs for Chagas disease (e.g. [5–7, 10]) have used the binary outcome of “sustained parasite negativity” as their primary efficacy endpoint. Although this endpoint has appealing simplicity and ease of interpretation, it ignores all quantitative assessments over time of parasite densities in blood, and does not take into account the number of samples taken. Thus, there is substantial variation in the overall sensitivity to detect parasitaemia during follow-up, resulting in imprecision in the end-point.

In this work we argue that the binary endpoint of “sustained negativity” versus “any positive PCR” is too crude for clinical trials of antiparasitic drugs in indeterminate Chagas disease. Given the extremely low blood parasite densities in these patients, a probabilistic assessment of cure based on the observed cycle threshold (Ct) values is more efficient for comparing different regimens or drugs. We present a detailed individual patient data meta-analysis focusing on the optimal methodology for interpreting serial quantitative PCR blood *T. cruzi* densities. By characterising the stochasticity in PCR readouts correctly at very low-density parasitaemias, we show that it is possible to derive more detailed information from clinical trial results in Chagas disease and thereby improve our understanding of optimal antiparasitic treatment.

## Material and Methods

### Clinical studies

#### Patient population

E1224 (enrolment between July 19, 2011, to July 26, 2012) and BENDITA (enrolment between Nov 30, 2016, and July 27, 2017) were randomised clinical trials conducted in outpatient units in Bolivia that specialised in Chagas disease. Both studies enrolled adults aged 18–50 years who had chronic indeterminate disease. Patients were eligible for enrolment in the study if they had a confirmed diagnosis of *T. cruzi* infection by qualitative PCR (at least one of three blood samples taken before randomisation) and positive conventional serology (a minimum of two positive tests; ELISA, recombinant ELISA, or indirect immunofluorescence); no acute or chronic health conditions and no signs or symptoms of the chronic cardiac or digestive form of Chagas disease. All patients provided written informed consent. Breastfeeding or pregnant women were excluded.

### Interventions studied

In both trials, investigators and sponsor staff were masked to treatment allocation and the randomisation list. In E1224, patients were partially blinded to treatment allocation; in BENDITA, patients were fully blinded to treatment allocation. In E1224, patients were randomised between five arms: benznidazole 300mg/day for 8 weeks (standard of care), fosravuconazole 400mg/week for 8 weeks; fosravuconazole 400mg/week for 4 weeks, fosravuconazole 200mg/week for 8 weeks, or placebo. The fosravuconazole allocation was placebo controlled, but the standard of care benznidazole regimen was open label. In BENDITA, patients were randomised between seven arms: benznidazole 300mg/day for 8 weeks (standard of care); benznidazole 300mg/day for 4 weeks; benznidazole 300mg/day for 2 weeks; benznidazole 150mg/day for 4 weeks; benznidazole 150mg/day for 4 weeks plus fosravuconazole 300mg/week for 8 weeks; benznidazole 300mg/week for 8 weeks plus fosravuconazole 300mg/week for 8 weeks, or placebo. Allocation was double-blind, double-dummy with matching placebos. In BENDITA, benznidazole was manufactured by ELEA (Abarax ELEA, 50mg and 100mg tablets), whereas in E1224, benznidazole was manufactured by Laboratorio do Estado de Pernambuco-LAFEPE (100mg tablets). In both trials, fosravuconazole was manufactured by EISAI Co. Ltd (100mg tablets) and was given in an initial 3-day loading dose, using the same daily dose as the subsequent weekly dose, or matching placebo. For example, the 300mg groups received 300mg on days 1, 2 and 3, and then 300mg weekly from weeks 2 to 8. Summary overviews of both trials are given in the Appendix, Figures S1 and S2.

### Follow-up

In E1224, blood sampling follow-up visits took place at one week (nominal day 7, one 5 ml blood sample), two weeks (nominal day 14, one 5 ml blood sample), and then at weeks 5, 9 and months 4, 6 and 12 (three 5 ml blood samples) [11]. In BENDITA, blood sampling follow-up visits took place on days 1, 2 and 3 (each patient had two visits randomly chosen from these 3 nominal days), and then days 7 and 14, and weeks 3, 5, 9, 11, and months 4, 6 and 12. At each visit three 5 ml blood samples were taken.

Both studies were implemented in accordance with Good Clinical Practice guidelines and the Declaration of Helsinki after approval by the ethical committees of the participating institutions (Universidad Mayor San Simón, CEADES, Hospital Clínic Barcelona, and ISGlobal). Both trials were pre-registered at ClinicalTrials.gov, numbers NCT01489228 and NCT03378661.

### PCR

The sampling schedule varied slightly across the two trials [6, 7]. At each visit between one and three 5 or 10ml venous blood samples were taken and each sample was then mixed with an equal volume of guanidine hydrochloride. DNA was extracted and between one and three 5uL aliquots were assayed for *T cruzi* by quantitative real-time PCR (qPCR) [11, 12]. Parasite load was estimated using a standard curve based on a *T. cruzi* stock from the discrete typing unit *T. cruzi V.* The PCR target was the *T. cruzi* satellite DNA, of which there are between 9000 and 35000 copies in each trypomastigote.

### Data collation

Clinical and laboratory data from both studies were shared with the Infectious Disease Data Observatory (IDDO) Chagas data platform for curation and standardisation, using the IDDO Study Data Tabulation Model (SDTM) Implementation Manual (https://www.iddo.org/tools-and-resources/data-tools). The curated and standardised data was pseudonymised individual participant data with no identifying variables, so approval was not required for the current analysis, as per the Oxford Tropical Research Ethics Committee requirements. The re-analysis was consistent with the primary intended purpose of the data collection so local ethics approval was not required. The research proposal was reviewed and approved by the DND*i* Scientific Advisory Committee.

### Meta-analysis

#### Analysis population

The primary analysis population for the efficacy analyses is a per-protocol population defined as individuals who took any active treatment for at least 80% of their allocated duration of active treatment. This choice of per protocol population is based on the observation that total dose is not the primary driver of efficacy, but total duration. The recorded treatment duration is defined as the interval of time between the first recorded dose and the last recorded dose. For example, if an individual randomised to 2 weeks benznidazole only took active treatment on days 1 and 12, this would be recorded as 12 days of treatment (i.e. 86% of total duration). Although a per-protocol analysis is potentially subject to post-randomisation confounding bias, this is unlikely as protocol deviations usually resulted from poor tolerability, which was unrelated to parasitological efficacy (the patients all had asymptomatic *T. cruzi* infections). As a sensitivity analysis, we estimated efficacy in the intention to treat population (ITT).

### Statistical model of efficacy

We modelled the serial qPCR values under a multi-level Bayesian regression model. This is a descriptive model of the serial qPCR data, which estimates the observed variability across time points, blood samples and PCR replicates (Figure 1). Trypomastigote blood densities were modelled on the Ct scale directly. Transformation from a Ct value to a number of parasites per ml is difficult because there is substantial variation in the number of sat-DNA target copies per parasite [13]. This copy number can vary across parasites within the same individual. Therefore, observed Ct values (with censoring at Ct = 40) are proxy measurements for the true unknown trypomastigote densities in each blood sample. Based on the data from the positive controls used for the qPCR in both studies (*T cruzi* V), a single parasite in a 5ml blood sample should theoretically correspond to an average Ct value between 38 and 40. Thus, in theory, mean Ct values up to 50 (assuming 2x multiplication per Ct) are plausible for an adult with a total blood volume of 5000 mL (approximately 2^10^ times lower: 1 parasite in 5000 ml of blood). For this reason, we allowed model estimated Ct values for each individual to be *≤* 50.

**Figure 1:**
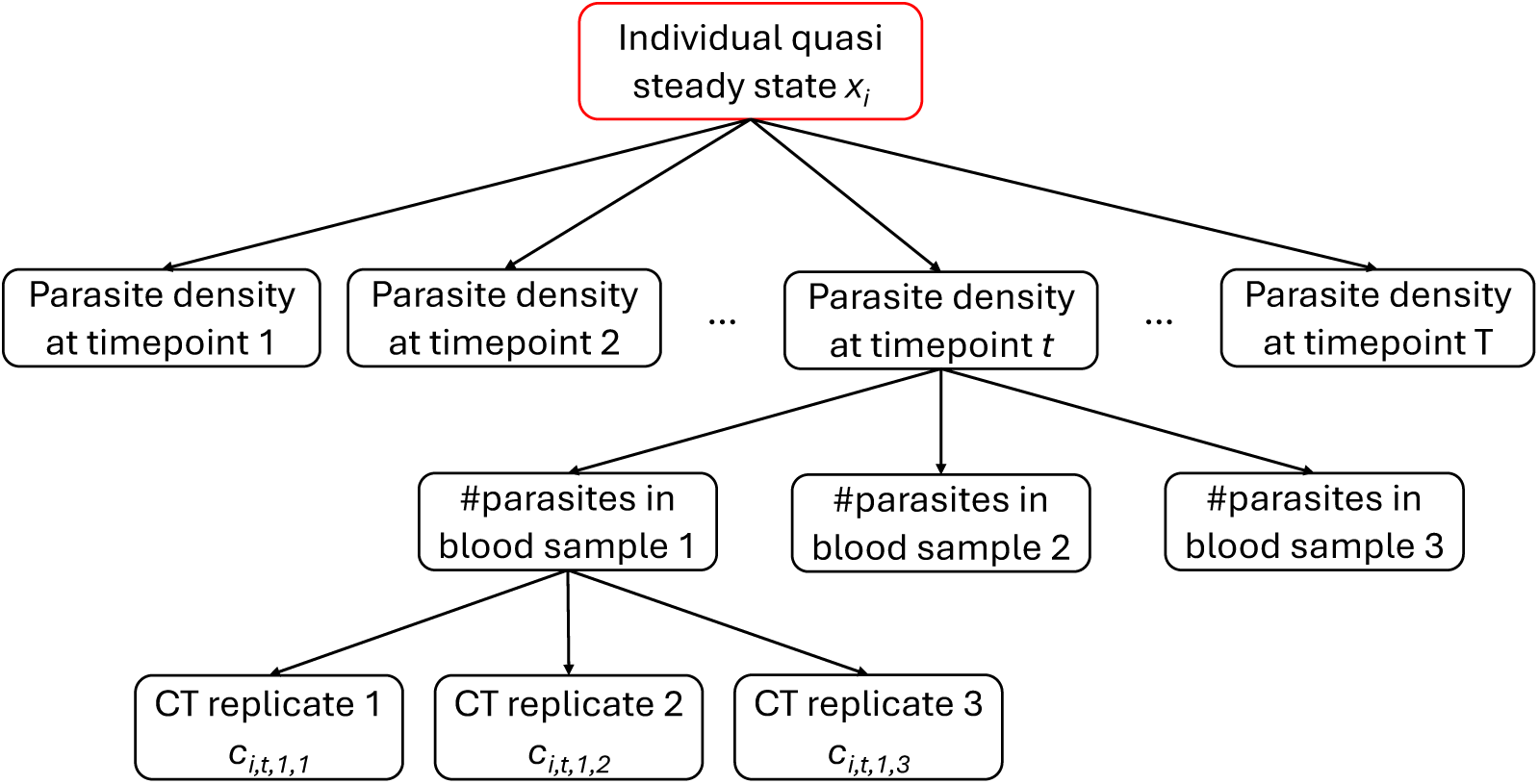
Graphical representation of the data structure for a given individual. We assume that individual *i* has a steady state trypomastigote density in blood denoted *x_i_* (treatment results in a step change in *x_i_* to either 0 (cure) or a lower positive value). Stochasticity in measurement of *x_i_* results from fluctuations over time, across blood samples, and the heteroskedastic PCR measurement error.

**Figure 2:**
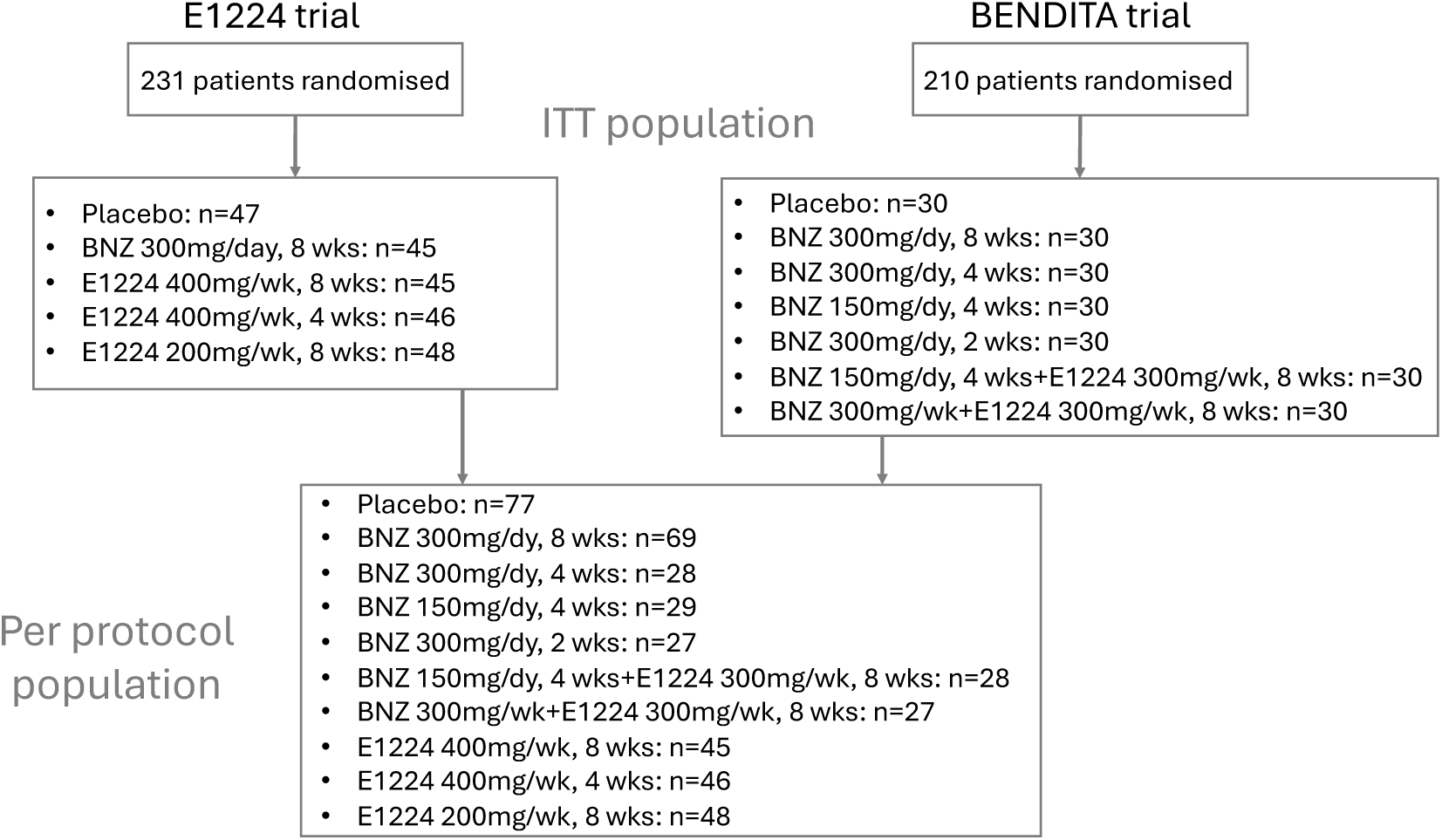
Consort diagram showing the number of patients randomised to each group (ITT population) and the number of patients allocated to placebo or who took active treatment for more than 80% of the allocated period (per protocol population).

The model assumes that each individual *i* is characterised by a steady state trypomastigote density in blood denoted *x_i_ ∈* (0, 50]. The steady state densities across individuals are assumed to be normally distributed, truncated at 50: *x_i_ ∼* Normal(*x*_pop_*, σ*_pop_). Each individual *i* has a total of *V_i_* study visits indexed as 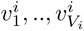. At each visit 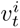, a total of 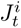 blood samples are taken (between 1 and 3). For each blood sample 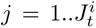, a total of 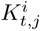 technical replicates are done. For most visits 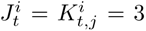 resulting in 9 observed Ct values per visit. We denote *c_i,t,j,k_ ∈* (0, 40] as the observed Ct value for individual *i* at visit *t* in blood sample *j* and technical replicate *k*. The observed Ct values *c_i,t,j,k_* are assumed to be normally distributed with censoring at 40 under a hierarchical model that incorporates nested variability by time (study visit), blood sample, and technical replicate, with normally distributed heteroskedastic error based on the ‘true’ blood sample Ct value *µ_i,t,j_*:

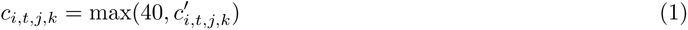

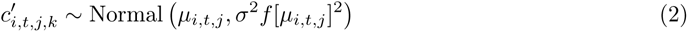

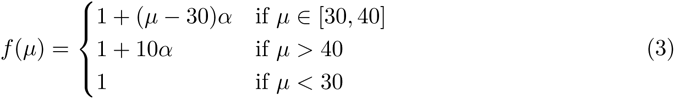

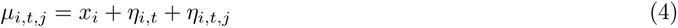

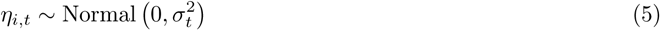

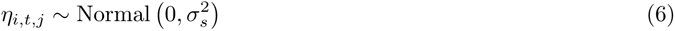

*µ_i,t,j_* is the mean trypomastigote density for individual *i* at time point *t* in blood sample *j*. This differs from the steady state *x_i_*by two additive random effect terms *η_i,t_* and *η_i,t,j_*. *η_i,t_* is the random effect by study visit (variance given by 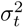), and *η_i,t,j_* is the random effect by blood sample (variance given by 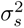). To ensure model identifiability, we assumed that *σ_s_ < σ_t_*. Instead of explicitly modelling the possibility that a blood sample by chance contains zero parasites, we allowed the random effect terms *η_i,t_, η_i,t,j_* to be unconstrained. This allows for model Ct values of greater than 50. Only the individual steady state values *x_i_* were constrained to be less than 50. The function *f* models the heteroskedastic variance as a function of the underlying expected Ct value. From graphical visualisation (Appendix Figure S4), this heteroskedasticity was apparent in the triplicate values for mean Ct values greater than 30, thus *f* was modelled as a piecewise linearly increasing function between 30 and 40. Because all data were censored at 40 it was not possible to know whether the stochasticity increased past 40 without strong assumptions, so we assumed constant variance for Ct values greater than 40.

Following treatment *T* (*i*), either the individual is cured with probability *p_T_* _(_*_i_*_)_ (no circulating trypomastigote parasites at any follow-up points) or they are not cured with probability 1 *− p_T_* _(_*_i_*_)_, but have a new lower steady state parasite density in blood. If the individual is cured, we assume 100% specificity of the PCR, i.e. P(*c_i,t,j,k_* = 40*|*cure) = 1 for all *t > t^∗^* where *t^∗^* is the end of treatment. If the individual is not cured, then:

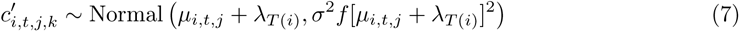

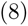

where *λ_T_*_(*i*)_ is the mean shift in steady state Ct values in those not cured.

##### Model assumptions

1. Each individual with *T. cruzi* infection has a steady state parasite density in blood greater than 1/5000 parasites per ml, represented on the Ct scale assumed to be less than 50 cycles.
2. Treatment with a given anti-parasitic drug either results in full cure, or lowers the average steady state density by a fixed amount.
3. Stochasticity in the measurement of parasite density in blood results from variation in time, blood sampling, and technical variation in the PCR output. These are nested independent random effects.
4. Variation in observed Ct values increases linearly as a function of the average Ct value (heteroskedastic noise).
5. Ct values *<* 40 are 100% specific (i.e. do not represent background fluorescence noise).

### Model fitting

We estimated the model parameters using Hamiltonian Monte Carlo, as implemented in *stan* [14]. The weakly informative prior distributions used for inference are given in the Appendix 6. The model likelihood for each individual was calculated as the marginal probability of the observed data under two possible scenarios (cured or not cured) using the log sum exp trick. We fitted a series of models to check validity and tested the model on simulated data. Four independent chains were run for 4,000 iterations. Model convergence and fitting were visually checked using the traceplots. R-hat values less than 1.01 were considered to indicate convergence. Parameter estimates are given as mean values and 90% credible intervals (CrI) defined as intervals between 5 and 95 percentiles of the posterior distributions.

### Tolerability and safety

Time to treatment discontinuation data was estimated from the ITT population using Kaplan-Meier survival analysis. We searched the Symptoms and Adverse Events (SA) domain for all terms related to dermatological, gastrointestinal, and neurological side effects, see Appendix 7 for exact terms used. For each category, the number of side effects were tabulated by individual and then compared across randomised treatment allocation under a zero-inflated Poisson regression model in the ITT population.

For liver toxicity, we estimated the effect of the daily dose and total duration of benznidazole, and the effect of the weekly dose of fosravuconazole on increases in alanine aminotransferase (ALT) relative to individual baseline values (mean of screening and day 0 values). All effects were modelled as additive. We did not include an effect for the duration of fosravuconazole treatment as this was completely co-linear with the lower 200mg/week regimen. ALT is the most specific of the liver enzymes measuring liver injury. ALT values were highly correlated with other measured “liver enzymes” (GGT, AST). We fit Bayesian penalised linear regression models using the package *rstanarm* to the log_2_ maximum observed ALT change from baseline between randomisation and week 11 (day 77) adjusted for age, sex and clinical site. For bone marrow toxicity, we fitted the same linear regression models to the observed minimum neutrophil count, adjusted for the baseline neutrophil count (mean of screening and day 0), with the same treatment effect terms.

### Code and data availability

All analysis code is openly accessible at https://github.com/jwatowatson/chagas-pcr-IPD-meta-analysis.

Data can be requested from the IDDO data sharing platform https://www.iddo.org/chagas/ data-sharing/accessing-data.

## Results

### Overview of study data

Clinical and laboratory data were obtained for a total of 441 Bolivian adult patients (E1224: *n* = 231; BENDITA: *n* = 210) of whom 73% were female (320 out of 441). BENDITA recruited most patients in the same clinical sites as the E1224 trial (Cochabamba and Tarija, 371 patients) as well as in an additional site in Sucre (70 patients). Clinical characteristics at enrolment were very similar across the two trials (Table 1). A total of 34,804 qPCR measurements were available from 5,402 unique time points spanning 449 patient years follow-up. The majority of patients adhered to their allocated treatment regimens; 90% of patients took at least 80% of their allocated active treatment. There was little loss to follow-up; 96% of patients had a follow-up visit at approximately one year after randomisation.

**Table 1:** Baseline characteristics of the per protocol population. Data show *n* (%) except for *^∗^* mean (standard deviation). BNZ: benznidazole; E1224: fosravuconazole.

|  | BNZ<br>150mg,<br>4wks | BNZ<br>150mg,<br>4wks<br>+<br>E1224<br>300mg,<br>8wks | BNZ<br>300mg,<br>2wks | BNZ<br>300mg,<br>4wks | BNZ<br>300mg,<br>8wks | BNZ<br>300mg/wk<br>+<br>E1224<br>300mg,<br>8wks | E1224<br>200mg,<br>8wks | E1224<br>400mg,<br>4wks | E1224<br>400mg,<br>8wks | Placebo |
| --- | --- | --- | --- | --- | --- | --- | --- | --- | --- | --- |
| N |  |  |  |  |  |  |  |  |  |  |
| Cochabamba | 9 (31.0) | 9 (32.1) | 5 (18.5) | 7 (25.0) | 31 (44.9) | 11 (40.7) | 24 (50.0) | 23 (50.0) | 23 (51.1) | 36 (46.8) |
| Sucre | 10 (34.5) | 8 (28.6) | 11 (40.7) | 13 (46.4) | 10 (14.5) | 6 (22.2) |  |  |  | 6 (7.8) |
| Tarija | 10 (34.5) | 11 (39.3) | 11 (40.7) | 8 (28.6) | 28 (40.6) | 10 (37.0) | 24 (50.0) | 23 (50.0) | 22 (48.9) | 35 (45.5) |
| Age<br>(years)* | 33.9 (8.3) | 30.8 (9.4) | 31.7 (7.5) | 33.2 (8.7) | 31.2 (8.8) | 33.9 (7.1) | 31.3 (8.8) | 27.8 (8.3) | 30.2 (8.9) | 31.9 (8.9) |
| Weight<br>(kg)* | 65.9 (7.7) | 64.8 (9.1) | 63.8 (8.3) | 64.0 (7.7) | 64.7 (10.9) | 66.5 (9.4) | 65.7 (9.5) | 58.7 (7.9) | 64.8 (11.4) | 62.8 (10.2) |
| Baseline<br>Ct* | 36.9 (2.3) | 37.5 (1.9) | 37.7 (1.6) | 36.9 (2.1) | 36.8 (2.6) | 36.0 (2.6) | 35.7 (3.1) | 36.6 (2.7) | 36.3 (2.9) | 36.7 (2.8) |
| Female | 19 (65.5) | 17 (60.7) | 20 (74.1) | 21 (75.0) | 44 (63.8) | 19 (70.4) | 37 (77.1) | 35 (76.1) | 32 (71.1) | 62 (80.5) |
| Male | 10 (34.5) | 11 (39.3) | 7 (25.9) | 7 (25.0) | 25 (36.2) | 8 (29.6) | 11 (22.9) | 11 (23.9) | 13 (28.9) | 15 (19.5) |

### Graphical visualisation of qPCR data

The heatmaps in Figures 3 and 4 show qPCR data from the two trials. Important points are evident from graphical visualisation of these data. First, there is considerable heterogeneity in qPCR read-out (i.e. Ct values) across triplicates done on the same blood sample, and across blood samples taken at the same time-point (Appendix Figure S4). Second, in the placebo treated groups in both trials, the observed distributions of Ct values remained fairly constant over time, although there was substantial inter-individual variability as noted previously [2]. Third, there is a very clear difference in parasite densities in blood following treatment between fosravuconazole and benznidazole. Fosravuconazole patients who were qPCR positive during follow-up (i.e. had recurrent parasitaemia) had Ct values similar to baseline, whereas the patients treated with benznidazole (apart from one individual who only received 4 days treatment) who were qPCR positive during follow-up had very low parasite densities and most had only a single positive PCR (out of 36 to 72 follow-up sample PCRs). In addition, there is evidence of a dose-response relationship for fosravuconazole (Figure 3), with a much greater effect for the 400mg dose taken over 8 weeks, although that effect was still much less than with any of the benznidazole regimens trialled.

**Figure 3:**
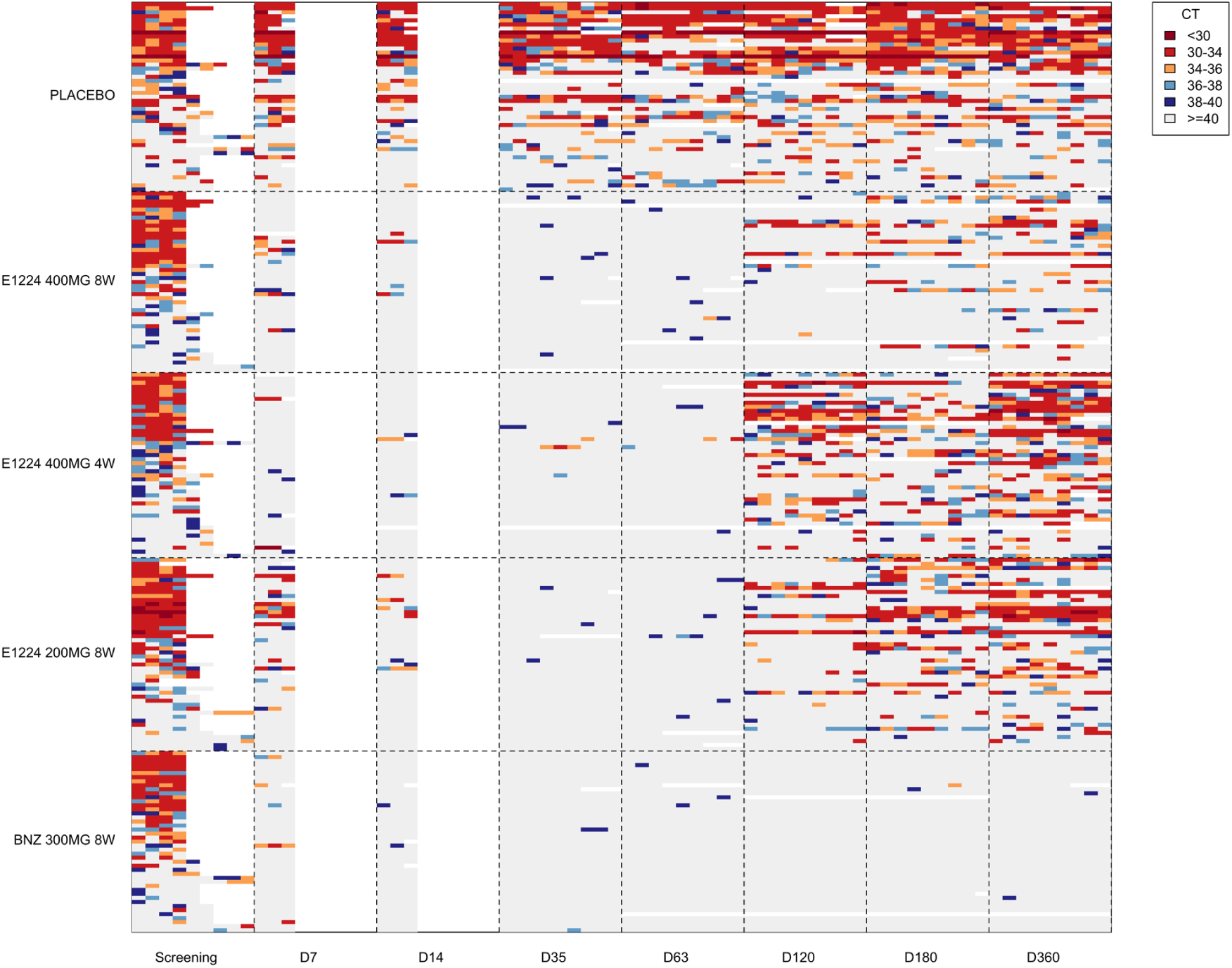
PCR data from the E1224 trial: *n* = 231 patients with 12,451 qPCR measurements (ITT population). Each row is one patient’s data, grouped by allocated treatment regimen and ordered within each group by mean Ct value at screening. At each time-point, up to 9 qPCRs were recorded (3 samples with up to 3 triplicates: 9 squares). Colours correspond to the recorded Ct value; white represents missing data, grey represents a Ct value of 40.

**Figure 4:**
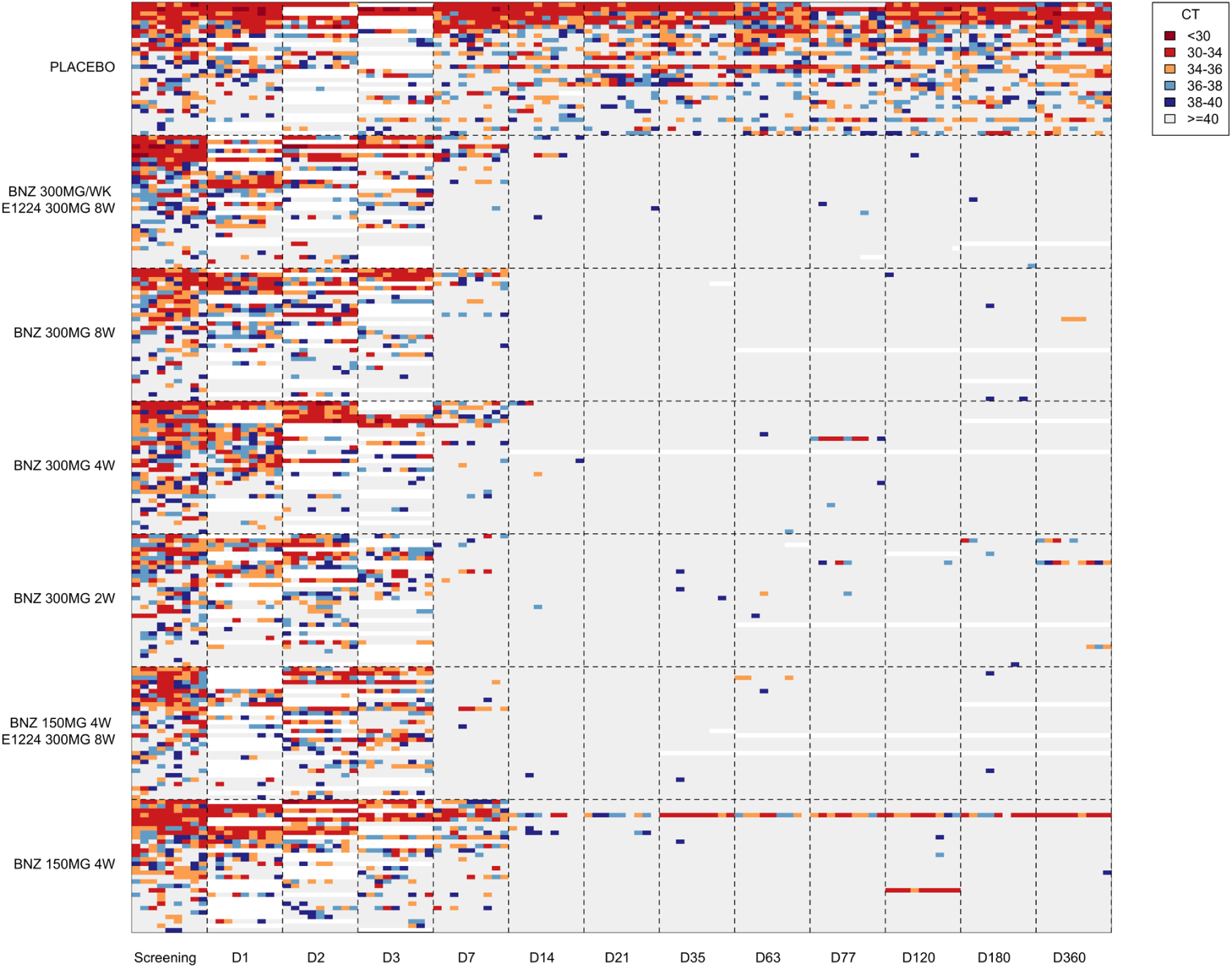
qPCR data from the BENDITA trial: *n* = 210 patients with 22,345 PCR measurements (ITT population). At each time-point, up to 9 qPCRs were recorded (3 samples with up to 3 triplicates: 9 squares). Colours as in Figure 3.

### Model estimated cure rates

We fitted the full statistical model to the serial qPCR data from patients randomised to placebo (*n*=77) or randomised to active treatment and who took any drug over a duration spanning at least 80% of the total allocated duration of active treatment (*n*=347). This included 14,748 Ct values, after excluding qPCR data from blood samples taken during treatment. This analysis excluded 17 individuals, one of whom only took 4 doses of 150mg of benznidazole over 6 days with no apparent effect on their parasite densities in blood. Under the model, spontaneous cure occurred in 4% of placebo treated individuals (95% CrI 1 to 9%; *n*=77). In comparison, an estimated 81% of individuals receiving 8 weeks of 300mg benznidazole daily were cured (95% CrI 70 to 89%, *n*=69). All the benznidazole regimens tested in the BENDITA trial, apart from the two-week regimen, had a similar estimated efficacy, with 95% CrI excluding cure rates less than 63% (Figure 5). The two-week benznidazole regimen was inferior to the standard 8-week regimen (posterior probability of lower cure rate was 0.95) with an estimated cure rate of 63% (95% CrI 43 to 81%). The estimated cure rates for the fosravuconazole arms were much lower but clearly indicated a dose-response relationship. Whereas the 200mg dose for 8 weeks had an estimated cure rate of 2% (95% CrI 0 to 11%), the 400mg dose for 4 weeks had an estimated cure rate of 9% (95% CrI 3 to 21%), and the 400mg dose for 8 weeks had an estimated cure rate of 23% (95% CrI 10 to 40%). A sensitivity analysis using the ITT population gave very similar estimates of cure and identical ranking of regimens (Appendix S5)

**Figure 5:**
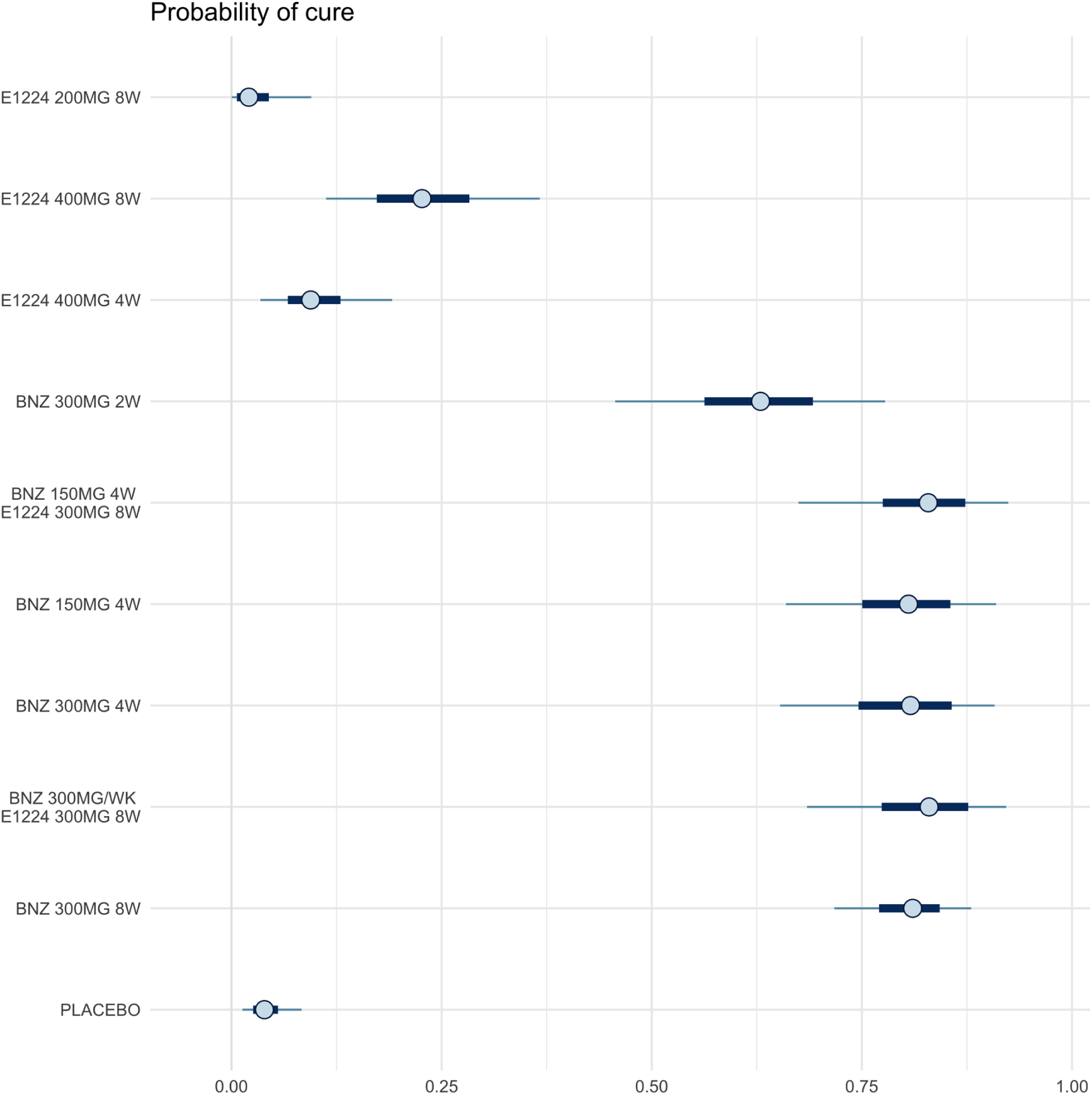
Estimated probabilities of cure in the per-protocol populations for the two trials (*n*=424). The median posterior estimates (circles) are shown along with 50% and 95% centred credible intervals. BNZ: benznidazole; E1224: fosravuconazole.

### Tolerability

In the ITT population, 24 patients discontinued treatment early, none of whom were in the placebo arm (Figure 6). The greatest number of discontinuations were in those allocated to benznidazole 300mg/day for 8 weeks (10/75, 13%). There were no treatment discontinuations in the two-week regimen, although this arm had the largest number of treatment interruptions (10/30, 33%). The intermittent 300mg benznidazole plus 300mg fosravuconazole weekly regimen had 4 treatment discontinuations (one because of neutropenia on day 21, and three because of raised liver enzymes). Two of these discontinuations affected administration of the final dose only.

**Figure 6:**
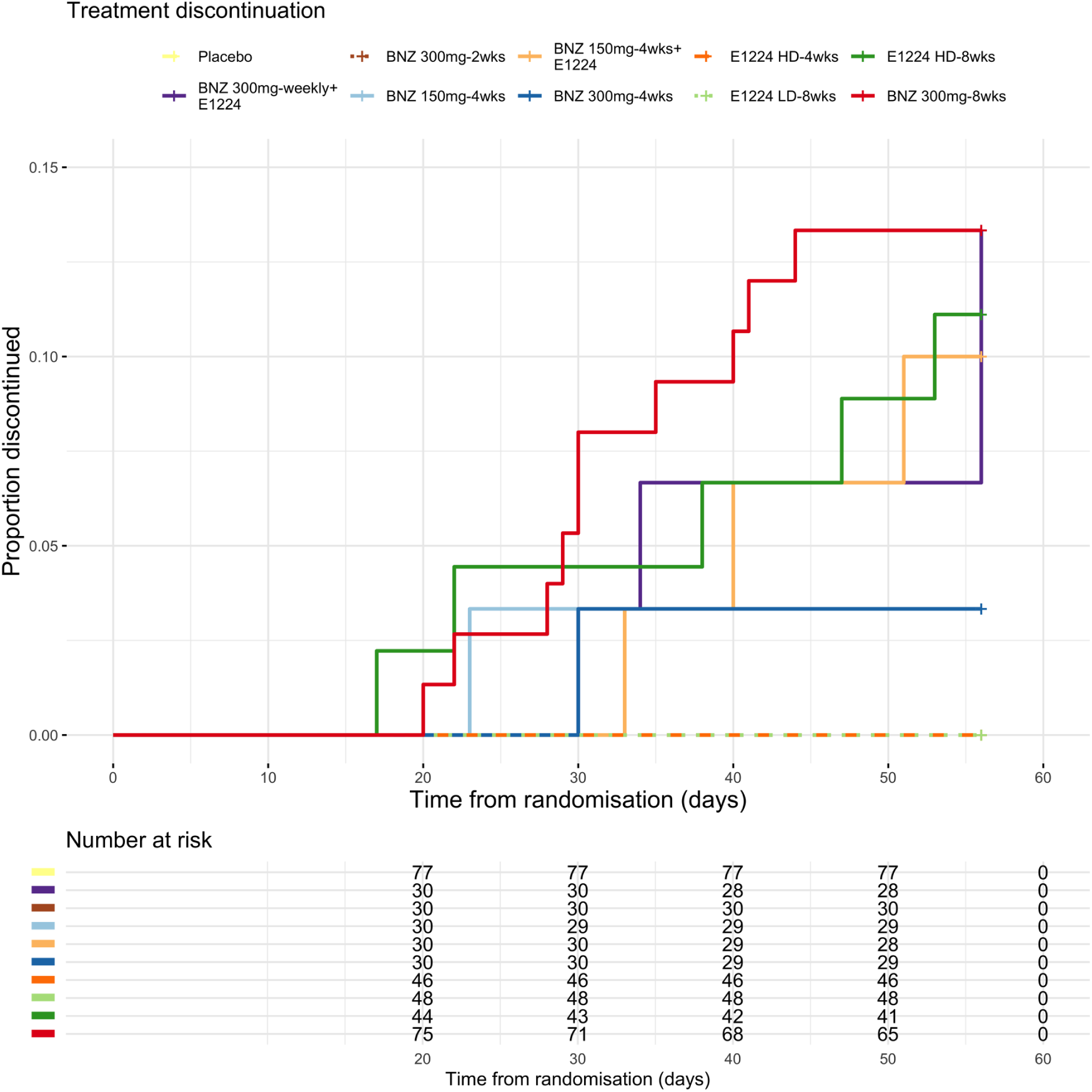
Kaplan-Meier curves showing the proportion of patients who discontinued treatment and time of discontinuation in the ITT population (*n*=441).

### Safety

There were no serious safety signals. Liver enzyme elevation is a known risk of treatment with fosravuconazole. Maximum log_2_ ALT changes relative to baseline were associated with male sex (15% lower, 95% CrI 5 to 25%), and the weekly fosravuconazole dose (3% higher per mg/kg increase, 95% CrI 0 to 5%), but not with age or the benznidazole dose or duration. Overall, 11 individuals had ALT rises greater than 8 times their baseline value, of which 5 were allocated to fosravuconazole 400mg weekly (*n*=45), 4 were allocated to fosravuconazole 300mg weekly (*n*=60) in combination with benznidazole, and 2 were allocated to benznidazole 300mg daily for 8 weeks (*n*=75). All cases of liver enzyme elevation were reversible on discontinuation of treatment and none met Hy’s law criteria.

Minimum observed neutrophil counts were associated with the duration of benznidazole treatment and the fosravuconazole weekly dose, but the estimated effects were very small (mean decrease of 56 neutrophils per uL per additional week of benznidazole treatment, 95% CrI 29 to 82; and a mean decrease of 28 per uL for each additional mg/kg weekly fosravuconazole dose, 95% CrI 10 to 46). Overall, 7 patients had a recorded neutrophil count *≤*1000 per uL, of whom one was in the placebo group; three were in the fosravuconazole monotherapy arms; one was in the 300mg benznidazole weekly + fosravuconazole arm; and two were in the benznidazole 8 weeks 300mg daily arm. In all cases, the neutropenia was transient, and none were associated with clinical infections.

## Discussion

The results of this individual patient data meta-analysis confirm that fosravuconazole alone is an inferior trypanocide to benznidazole, even at the higher dose of 400mg/week for 8 weeks. Although the sample sizes are too small to give precise estimates of cure, the 300mg/week benznidazole regimen given for eight weeks (with fosravuconazole) and the four-week regimens (irrespective of dose) performed as well as the current standard eight-weeks regimen of daily administration. In contrast, the estimated parasite recurrence rate with the two-week benznidazole regimen was approximately twice that following the four-week or eight-week regimens. The clinical significance of this difference is uncertain, but taken together, these results suggest that the current eight-week 300mg/day benznidazole regimen is excessive and provide further support for the evaluation of weekly administration and for shorter courses of benznidazole [5, 7, 15–18]. Shorter or intermittent treatment regimens bring enormous advantages including better adherence, better tolerability, greater safety and lower cost. Comparison of tolerability and safety of the intermittent regimen with the daily regimens of benznidazole in this study is complicated because the intermittent regimen also included fosravuconazole 300mg/week over 8 weeks. Treatment discontinuations in this arm (particularly the raised ALT levels) are likely to be due to fosravuconazole and not benznidazole.

A major contribution of this work is the proposed statistical methodology for the analysis of serial qPCR data. Although the methodology is imperfect, it provides a coherent model framework for ranking the efficacy of anti-trypanosomal drug regimens. The model takes into account the variable number of follow-up visits, missing data, the variable number of PCR replicates, and the low sensitivity of PCR to detect residual parasites in blood. Even though the model is misspecified, the ranking of the interventions is highly robust. Our results have several implications for the conduct of therapeutic assessments in Chagas disease. Quantification of the therapeutic efficacy of active drugs is very sensitive to the specificity of qPCR results when the parasite density is close to the limit of accurate quantification. Larger sample sizes are needed to characterise therapeutic responses more accurately and more baseline assessment (i.e. > 1 baseline qPCR positive sample) is needed to evaluate the pre-treatment parasitaemia. Selecting individual volunteers for clinical trials based on a single “positive” qPCR result will include peak values which revert to lower average steady state densities (“reversion to the mean”). Individuals with very low baseline values will contribute very little information regarding drug efficacy. The significance of a single weakly “positive” qPCR following treatment among a triplicate of samples (i.e. in which the other two samples are “negative”) is uncertain. The likelihood that such a result is a genuine positive depends on many factors including the Ct value itself, the results of other tests in the temporal series, the performance of the qPCR assay, the drug used and the exposure to the treatment (a function of duration, dosage and individual pharmacokinetics). A probabilistic approach is therefore required to interpret individual patient data in the context of clinical trials.

The therapeutic objective in treating chronic indeterminate Chagas disease is to prevent the long-term pathological consequences of the chronic parasitic infection, which mainly affect the heart and the gastrointestinal tract. These occur decades after initial infection, so shorter term assessments are needed to evaluate and compare parasitological outcomes of anti-trypanosomal treatments. But assessment of parasitological cure in chronic indeterminate Chagas disease is challenging. Despite the development and validation of remarkably sensitive qPCR methods to quantitate low density parasitaemias, most quasi steady state *T. cruzi* parasite densities are either below or close to the limits of qPCR detection (1 parasite in 5-15mL of blood). Substantial efforts have been made to standardise and harmonise qPCR assessment in Chagas disease and support clinical trials [12, 19], but the interpretation of the trial results has been uncertain. As in other microbe detection assays, most qPCR assays are terminated after 38 to 40 cycles as the enzymes lose activity and serial PCR amplification may eventually yield a non-specific false “positive” result. However, the majority of post treatment parasite densities in chronic Chagas disease give qPCR signals very close to this cut-off value. PCR readout is inherently more variable after 35 cycles (heteroskedasticity). The resulting stochasticity, compounded by temporal parasite density fluctuations, therefore results in varying PCR “positivity” rates over time. The BENDITA trial qPCR results show that benznidazole reduces circulating parasite densities rapidly, and may result either in “cure” (i.e. sustained qPCR negativity), or a new lower quasi-steady state density in which there is a substantially lower probability of detecting parasitaemia. This assumes that the observed Ct values close to 40 do not represent false positive results. The pathological consequences of a substantially lower steady state parasitaemia and its longer-term evolution are uncertain, but, should reflect lower cumulative tissue burdens and thus a reduced risk of subsequent organ dysfunction. Distinguishing full cure from a lower steady state is difficult and necessitates taking multiple blood samples to improve sensitivity (Appendix Figure S6). Because of low sensitivity, infrequent sampling will variably underestimate the true proportion of patients with recurrent parasitaemia.

#### Optimal trial design considerations for phase 2 studies of antiparasitic drugs for Chagas disease

1. *Selection of patients*: there should be a higher threshold of PCR positivity for inclusion into the trial. Patients who are only weakly positive at a single PCR replicate (out of 9) will likely have very low parasite densities and thus will contribute very little information regarding treatment efficacy.
2. *During treatment* : both efficacious (eg benznidazole) and inefficacious treatments (fosravuconazole) clear parasites quickly from blood. Blood PCR data taken during treatment are not useful for interpreting the outcomes.
3. *After treatment* : the total number of blood samples taken will determine the overall sensitivity in detecting circulating parasites in the blood. As recurrences often reach detectable parasitaemias many months after receiving treatment, these blood samples should be taken later rather than sooner after treatment, over a period of 1-2 years. In theory, 13 independent 5ml blood samples can detect an average parasite density of 0.01 parasites per ml (50 trypomastigotes in the entire blood volume of an adult), Figure S3.

There are several important limitations to this meta-analysis of the E1224 and BENDITA trials. First, the model estimates of cure are critically dependent on the assumption that Ct values less than 40 do not represent false positives. Of the 41 patients treated with benznidazole who were PCR positive during follow-up, 31 were positive only on a single PCR, for which 24 (i.e. over half) had Ct values *≥* 38. If these are false positives then the true treatment efficacies of all benznidazole regimens are higher than our estimates, although the ranking of relative efficacies would not change. Clinical trials of Chagas treatment using PCR as their primary endpoint should have pre-specified protocols in place to verify that results are not driven by false positives. Single positive PCR results should have new sets of technical replicates re-done for verification, preferably using a different PCR target (for example kinetoplastid DNA). Second, the estimates of cure are currently based on one year follow-up only. It is far from certain that one year follow-up is sufficient. The data from the fosravuconazole treated patients indicate a temporal trend between visits at 4 and 6 months, which presumably reflects slow intra-host parasite population re-growth. Whether this is also the case for more potent treatments, such as benznidazole, remains to be seen. Third, the design of the BENDITA trial was inefficient. BENDITA evaluated benznidazole regimens of shorter duration, lower daily dose, intermittent dosing, and in combination with fosravuconazole. This represents a 4-dimensional exposure-response evaluation. The most efficient approach to evaluation would have been a factorial design, which is the only design that can disentangle the contributions of each dimension. Because BENDITA was not factorially randomised, we cannot estimate the contribution of fosravuconazole in the intermittent regimen. However, given the very low efficacy of 400mg/week fosravuconazole monotherapy, this is unlikely to be substantial. In addition, the precision of the estimates is very low for most treatment groups, notably the two weekly regimen and the intermittent regimen. This is of particular importance given the variability associated with low density *T. cruzi* parasitaemias. Finally, the clinical manifestations and drug susceptibility of *T. cruzi* infections vary widely. The results with the *T. cruzi* group 5 discrete typing unit (DTU) infections prevalent in Bolivia may not be directly applicable to other regions where different DTUs are prevalent.

These clinical studies support earlier investigations in Chagas disease both in animal models and in humans, which suggest that the currently recommended 8 weeks daily dosing benznidazole regimen is excessive [15, 17, 20, 21]. Weekly dosing for eight weeks or daily dosing over four weeks both appear effective in people from Bolivia with *T. cruzi* infection, and both are less expensive and less toxic than the current regimen, thus increasing access to these treatments. Large factorially randomised trials are needed to determine the contributions of duration, dose and intermittency to benznidazole efficacy.

## Data Availability

Data can be requested from the IDDO data sharing platform https://www.iddo.org/chagas/data-sharing/accessing-data.

## Acknowledgements

JAW is a Sir Henry Dale Fellow funded by the Wellcome Trust (223253/Z/21/Z). NJW is a Principal Research Fellow funded by the Wellcome Trust (093956/Z/10/C). This research is funded by the Wellcome Trust grant “Optimising pharmacometric assessment in phase 2 studies of Chagas disease” (222754/Z/21/Z) awarded to JAW, CC, SSE, PJG, and NJW.

DND*i* received financial support to conduct these clinical studies from the following donors: Strategic Translation Award from the Wellcome Trust (grant number 095422); Global Health Innovative Technology Fund, Japan; Department for International Development (UK Aid), UK; German Federal Ministry of Education and Research through Kreditanstalt für Wiederaufbau, Germany; Dutch Ministry of Foreign Affairs, Netherlands; Brazilian Ministry of Health, Brazil; Associação Bem-Te-Vi Diversidade, Brazil; Fundação Oswaldo Cruz (Fiocruz), Brazil; Starr International Foundation, Switzerland. Ministry of Foreign Affairs, Spain; Rockefeller Foundation, USA; and anonymous donors. The Platform for a Comprehensive Care of Patients with Chagas disease in Bolivia is a collaborative project between CEADES of Health and Environment; Universidad Mayor de San Simon in Cochabamba, Bolivia; Juan Misael University Saracho, Tarija, Bolivia; and ISGlobal (Barcelona Institute for Global Health, Spain), and supported by the National Chagas Control Program in Bolivia. The Platform is funded by the Spanish Agency for Cooperation and Development (grant number 10-CO1-039). For its overall mission, DND*i* also receives funding from the Swiss Agency for Development and Cooperation, Switzerland and Médecins Sans Frontières.

This research was partly funded by Wellcome. A CC BY or equivalent licence is applied to the author accepted manuscript arising from this submission, in accordance with the grant’s open access conditions.

## 1 Trial Design

**Figure S1:**
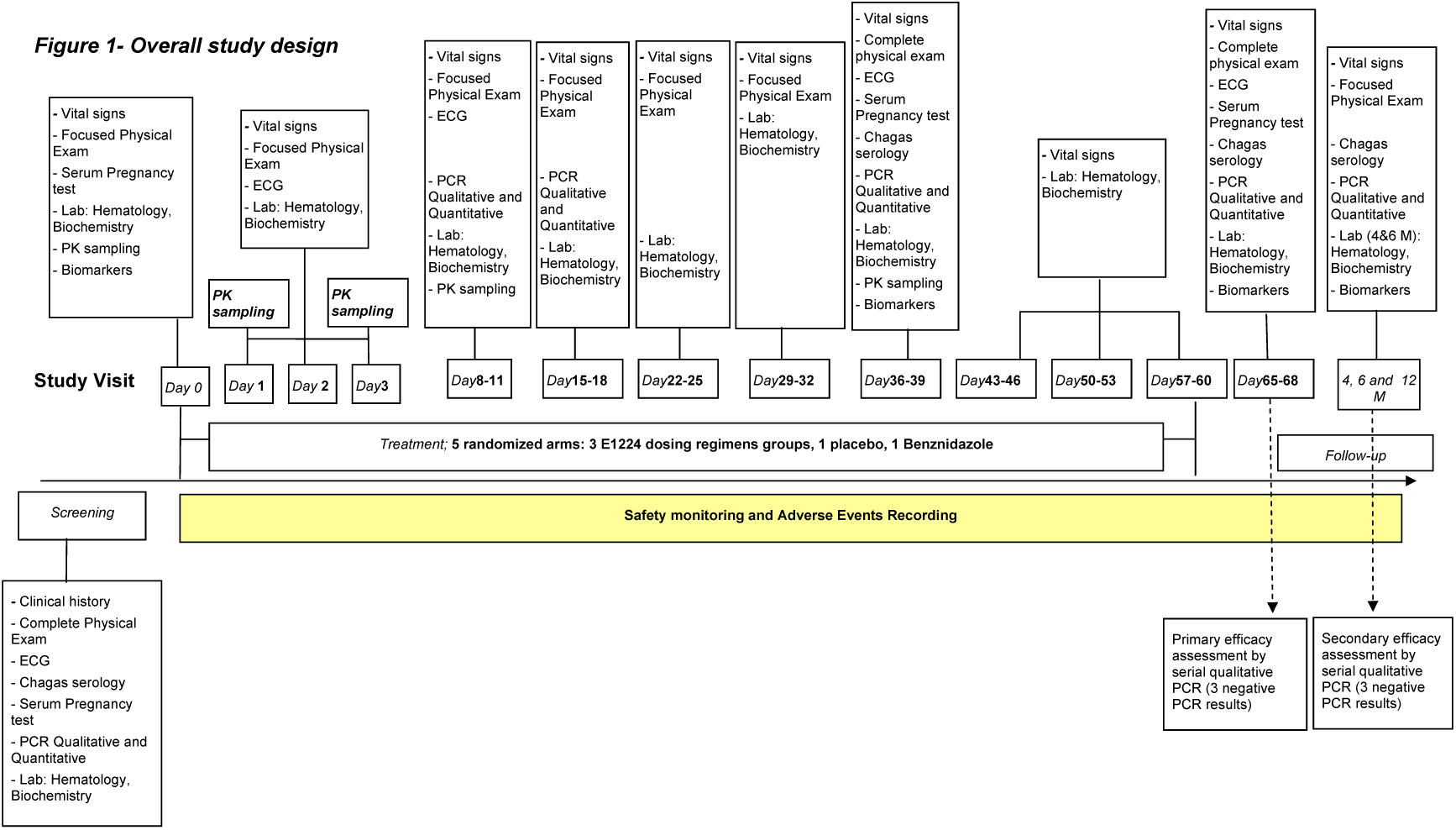
Design of the E1224 trial [6]. Taken from the trial protocol.

**Figure S2:**
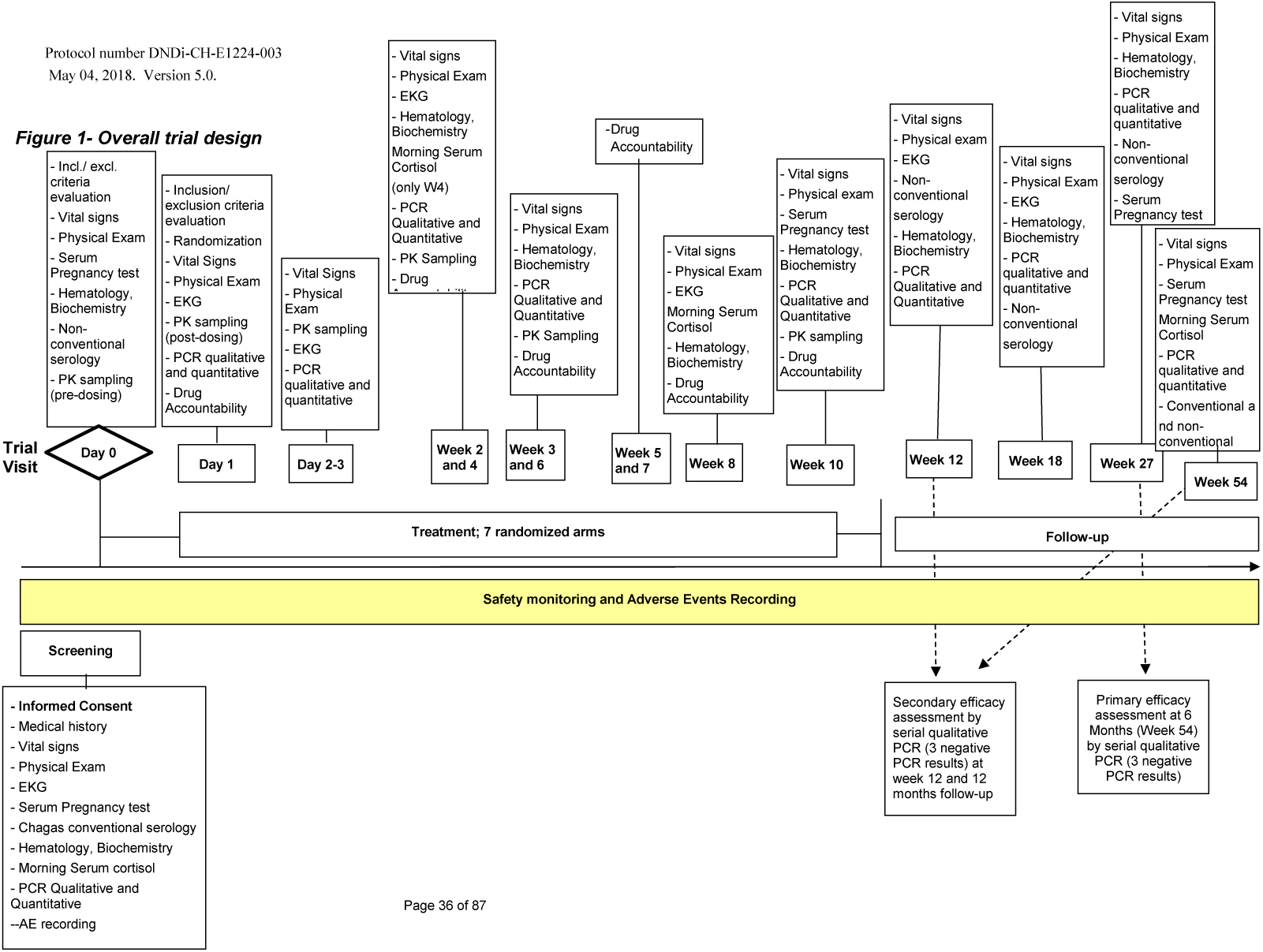
Design of the BENDITA trial [7]. Taken from the trial protocol.

## 2 Optimal design considerations

**Figure S3:**
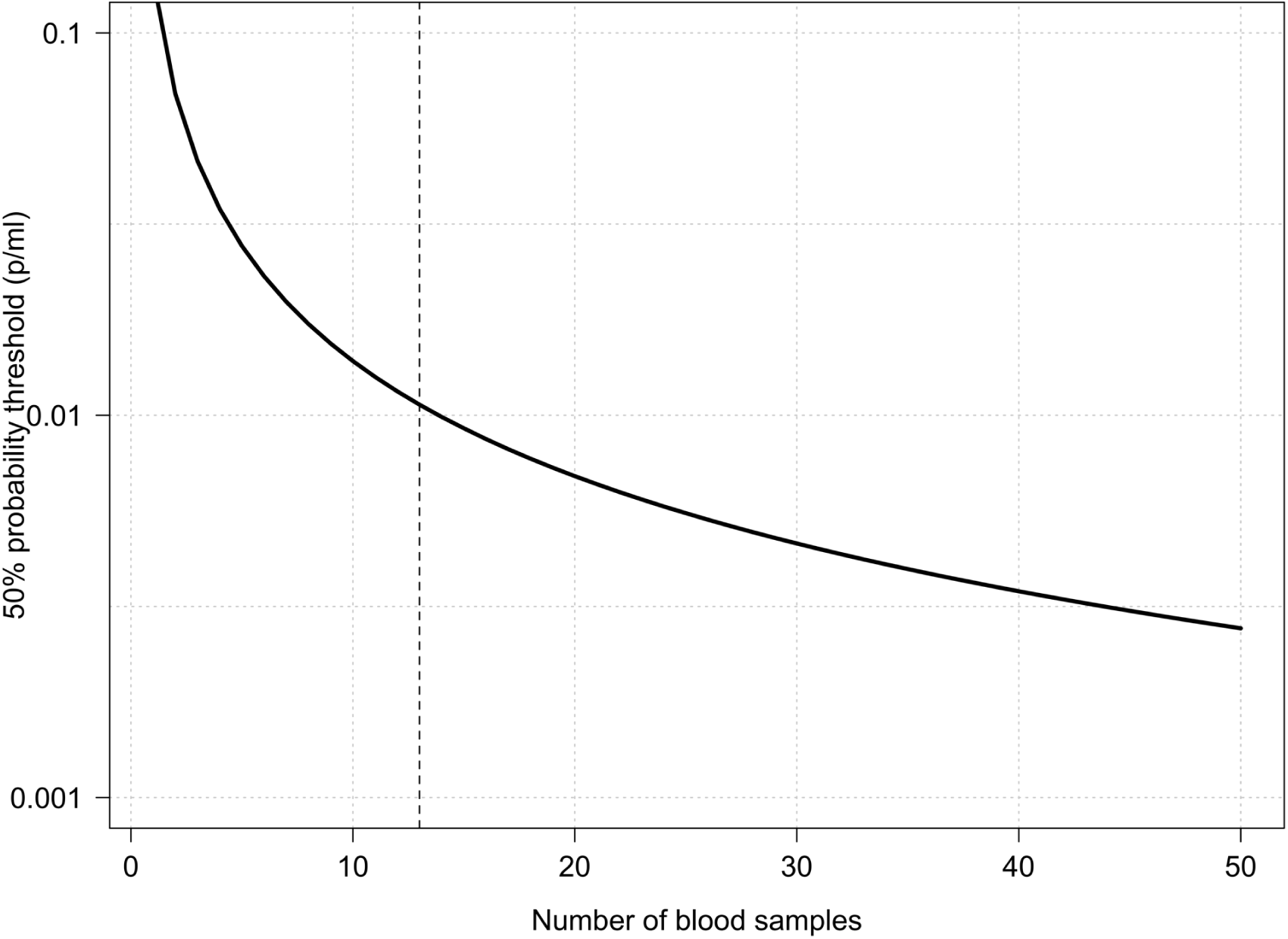
Theoretical sensitivity of sampling at least one parasite in a 5ml blood sample as a function of the number of follow-up samples taken. 13 samples results in a 50% chance of sampling at least one parasite for a mean parasite density of 0.01 per ml (ie 50 parasites in 5000 ml of blood).

## 3 Heteroskedasticity in PCR readout

**Figure S4:**
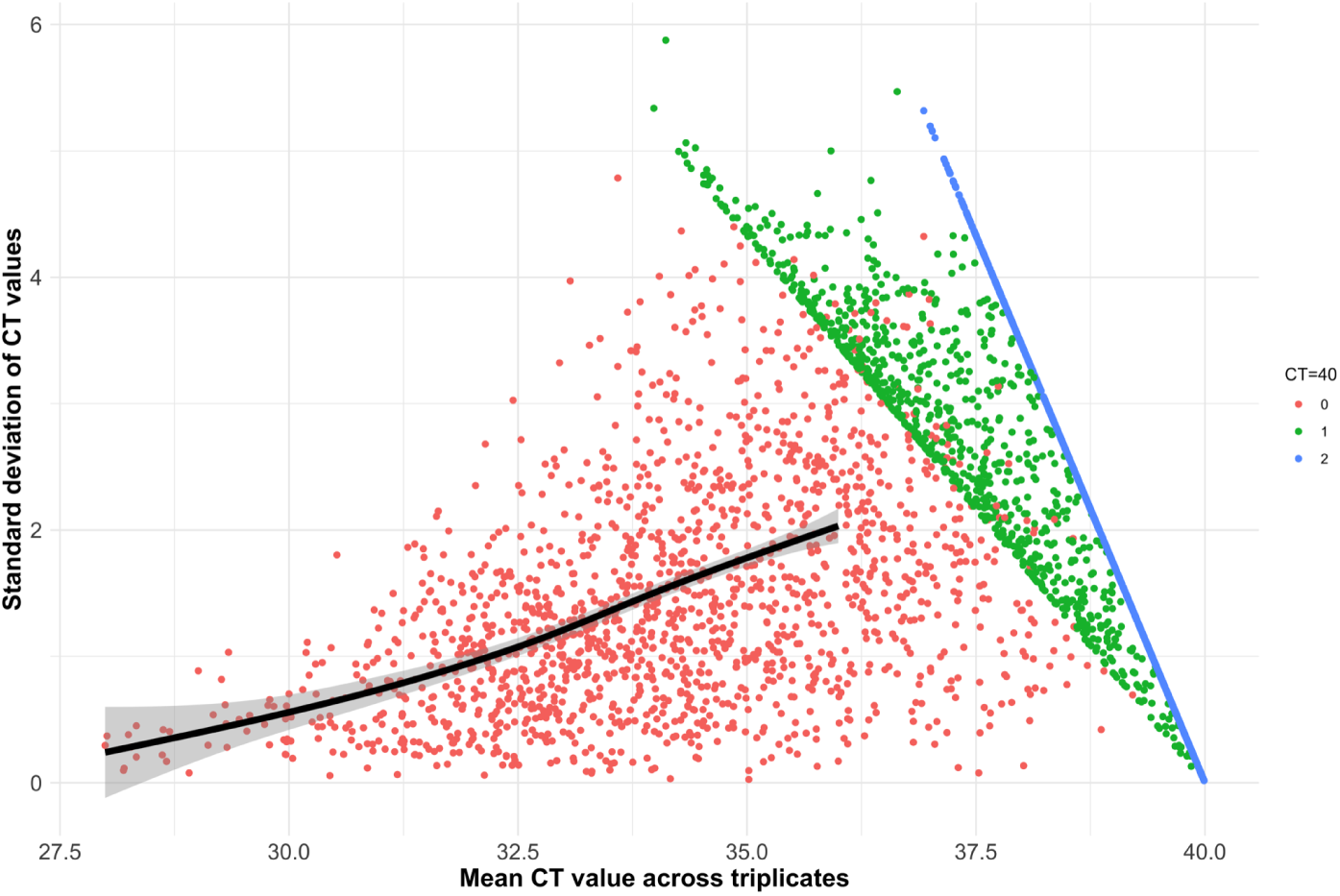
The relationship between the mean value and the standard deviation among all samples for which qPCR was done in triplicate and where at least one sample had a Ct value *<* 40 (*n*=3,482 samples). The points are coloured by the number of technical replicates with a Ct value *<* 40. Red: all Ct values *<* 40; green 2/3 Ct values *<* 40; blue: 1/3 Ct values *<* 40. The black line shows a spline fit to the data with all three triplicates *<* 40 (red points) with mean value between 27 and 36 (above 36 the spline fit is biased downwards).

## 4 ITT efficacy analysis

**Figure S5:**
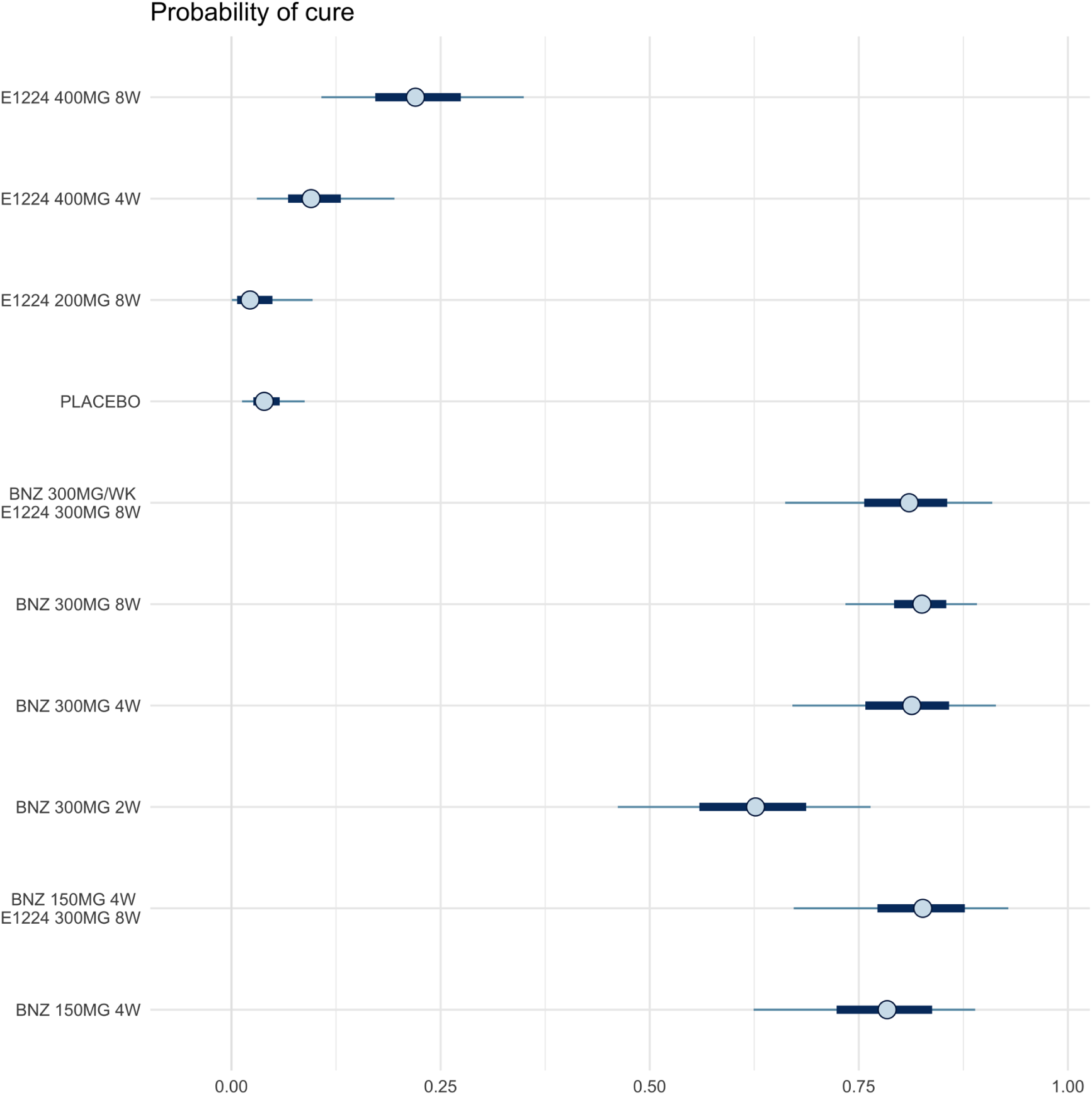
Estimated probabilities of cure in the ITT population (*n*=441). BNZ: benznidazole; E1224: fosravuconazole.

## 5 Theoretical sensitivity

Figure S6 shows the probability that at least one blood sample of 5ml contains at least one *T. cruzi* parasite as a function of the parasite density in blood and the number of blood samples taken. If we assume that each blood sample is independent across time-points, then 5 visits with three 5ml samples taken at each visit result in 15 independent draws. This would provide a greater than 50% chance that one of these 15 samples would contain at least 1 parasite for densities as low as 0.01 parasites per ml (i.e. 50 trypanosomes circulating in an adult with 5000ml of blood). There are only small gains in increasing the number of samples beyond 15. However, it may be that temporal fluctuations in blood densities mean that the number of parasites per sample are correlated for a given visit. This would imply that fewer samples taken per visit, but more follow-up visits (especially over a longer time period) would increase the chance of detecting parasites in blood. It also emphasises the importance of increasing PCR sensitivity: i.e. being able to detect reliably 1 parasite in a 5ml blood sample.

**Figure S6:**
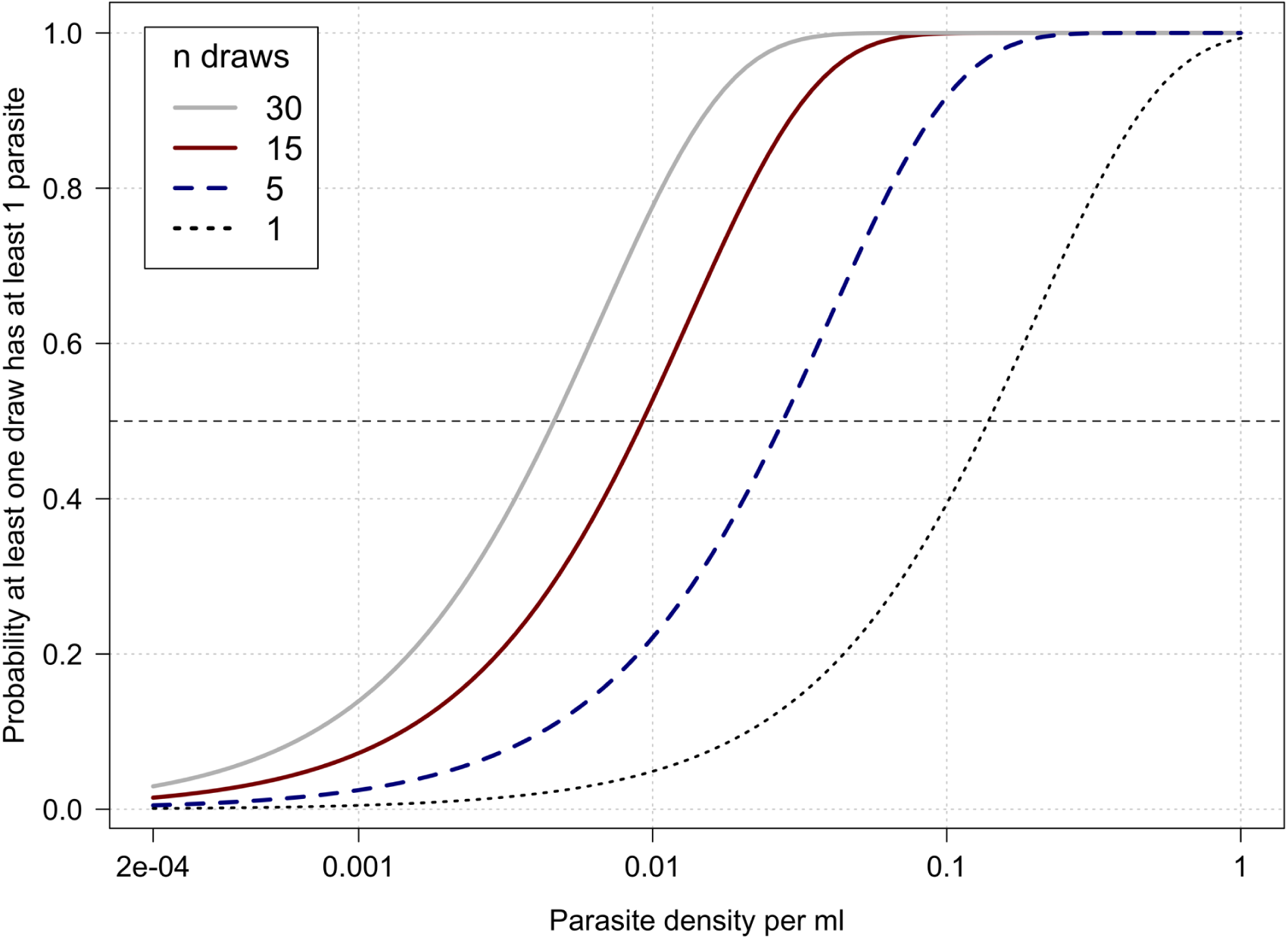
Probability of sampling at least one parasite in one 5ml blood sample as a function of the parasite density in blood and the number of independent blood samples taken. This assumes a Poisson distributed number of parasites per blood sample (mean given by the blood density mean times 5).

## 6 Prior distributions

Prior distributions were specified as follows:

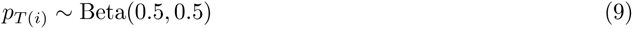

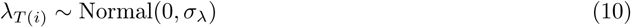

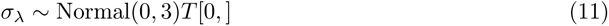

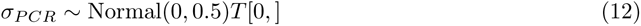

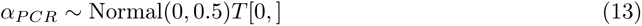

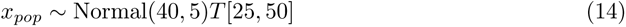

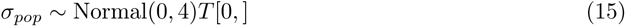

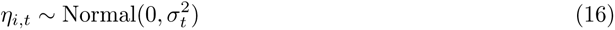

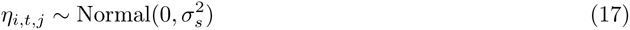

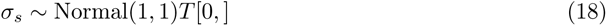

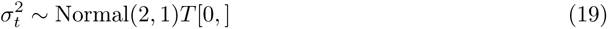

## 7 Tolerability definitions

We defined three categories of adverse events: skin, neurological and gastrointestinal. The number of events was calculated as the number of unique records with terms matching specific search strings.

For skin related disorders we selected all recorded SA events that included in the SATERM variable: erythem* OR papules OR prurit* OR itch* OR dermatit* OR rash OR urtic* OR skin OR dermat* OR desquamat* OR scratch*; or defined in the SADECOD variable as: Itching OR Urticaria OR Eruption of skin.

For gastrointestinal side effects we selected all events that included in the SATERM variable: eating OR stool OR epigastralgia OR hiatal hernia OR vomit* OR flatulence OR abdominal OR epigastric OR pyrosis OR stomach OR gastri OR reflux OR nause* OR constip OR diarrhea OR intestinal OR dyspepsia OR meteorism OR pirosis; or defined in the SADECOD variable as: Abdominal pain OR Diarrhea OR Constipation OR Stomach ache.

For neurological side effects we selected all events that included in the SATERM variable: cognition OR headache OR dizz* OR vertig OR migrain OR parasthesia OR paresthe* OR peripheral OR dysestesia OR hypoesthes OR neuropathy OR headburn OR somnolence OR hypersomnia OR neuritis OR facial paralysis OR lipothymia OR neuro* OR seizures; or defined in the SADECOD variable as: headache OR Dizziness OR Seizure OR Disorder of vision OR Insomnia OR Anxiety.

